# Sample-Efficient Deep Learning for COVID-19 Diagnosis Based on CT Scans

**DOI:** 10.1101/2020.04.13.20063941

**Authors:** Xuehai He, Xingyi Yang, Shanghang Zhang, Jinyu Zhao, Yichen Zhang, Eric Xing, Pengtao Xie

## Abstract

Coronavirus disease 2019 (COVID-19) has infected more than 1.3 million individuals all over the world and caused more than 106,000 deaths. One major hurdle in controlling the spreading of this disease is the inefficiency and shortage of medical tests. There have been increasing efforts on developing deep learning methods to diagnose COVID-19 based on CT scans. However, these works are difficult to reproduce and adopt since the CT data used in their studies are not publicly available. Besides, these works require a large number of CTs to train accurate diagnosis models, which are difficult to obtain. In this paper, we aim to address these two problems. We build a publicly-available dataset containing hundreds of CT scans positive for COVID-19 and develop sample-efficient deep learning methods that can achieve high diagnosis accuracy of COVID-19 from CT scans even when the number of training CT images are limited. Specifically, we propose a Self-Trans approach, which synergistically integrates contrastive self-supervised learning with transfer learning to learn powerful and unbiased feature representations for reducing the risk of overfitting. Extensive experiments demonstrate the superior performance of our proposed Self-Trans approach compared with several state-of-the-art baselines. Our approach achieves an F1 of 0.85 and an AUC of 0.94 in diagnosing COVID-19 from CT scans, even though the number of training CTs is just a few hundred.

## I. Introduction

**C**oronavirus disease 2019 (COVID-19) is an infectious disease that has infected more than 1.3 million individuals all over the world and caused more than 106,000 deaths^1^, as of April 11 in 2020. One major hurdle in controlling the spreading of this disease is the inefficiency and shortage of tests. The current tests are mostly based on reverse transcription polymerase chain reaction (RT-PCR). It takes 4-6 hours to obtain results, which is a long time compared with the rapid spreading rate of COVID-19. Besides inefficiency, RT-PCR test kits are in huge shortage. As a result, many infected cases cannot be timely identified and continue to infect others unconsciously.

To mitigate the inefficiency and shortage of existing tests for COVID-19, many efforts have been devoted to searching for alternative testing methods. Several studies [1] have shown that computed tomography (CT) scans manifest clear radiological findings of COVID-19 patients and are promising in serving as a more efficient and accessible testing manner due to the wide availability of CT devices that can generate results at a fast speed. Further, to alleviate the burden of medical professionals in reading CT scans, several works [2] have developed deep learning methods that can automatically interpret CT images and predict whether the CTs are positive for COVID-19. While these works have shown promising results, they have two limitations. First, the CT scan datasets used in these works are not sharable to the public due to privacy concerns. Consequently, their results cannot be reproduced and the trained models cannot be used in other hospitals. Besides, the lack of open-sourced annotated COVID-19 CT dataset greatly hinders the research and development of more advanced AI methods for more accurate CT-based testing of COVID-19. Second, these works require a large collection of CTs during model training to achieve performance that meets the clinical standard. Such a requirement is stringent in practice and may not be met by many hospitals, especially under the circumstances that medical professionals are highly occupied by taking care of COVID-19 patients and are unlikely to have time to collect and annotate a large number of COVID-19 CT scans.

In this work, we aim to address these two problems by (1) building a publicly-available dataset containing hundreds of CT scans that are positive for COVID-19 and (2) developing sample-efficient deep learning methods that can achieve high diagnosis accuracy of COVID-19 from CT scans even when the number of training CT images are limited. We first collect the **COVID19-CT** dataset, which contains 349 CT images with clinical findings of 216 COVID-19 patient cases. The images are collected from medRxiv and bioRxiv papers about COVID-19. CTs containing COVID-19 abnormalities are selected by reading the figure captions in the papers. We manually remove artifacts in the original images, such as texts, numbers, arrows, etc. Figure 1 shows some examples of the COVID-19 CT scans. To our best knowledge, it is the largest COVID-19 CT dataset to date. And all the images are open to the public for research purpose. Given this dataset, we develop deep learning (DL) methods to perform CT-based diagnosis of COVID-19. Though largest among its kind, COVID19-CT is still limited in image number. DL models are data-hungry, which have high risk of overfitting when trained on small-sized dataset. To address this problem, we develop sample-efficient methods to train highly-performant DL model in spite of data deficiency. Specifically, we investigate two paradigms of learning approaches for mitigating data deficiency: transfer learning and self-supervised learning.

**Fig. 1.**
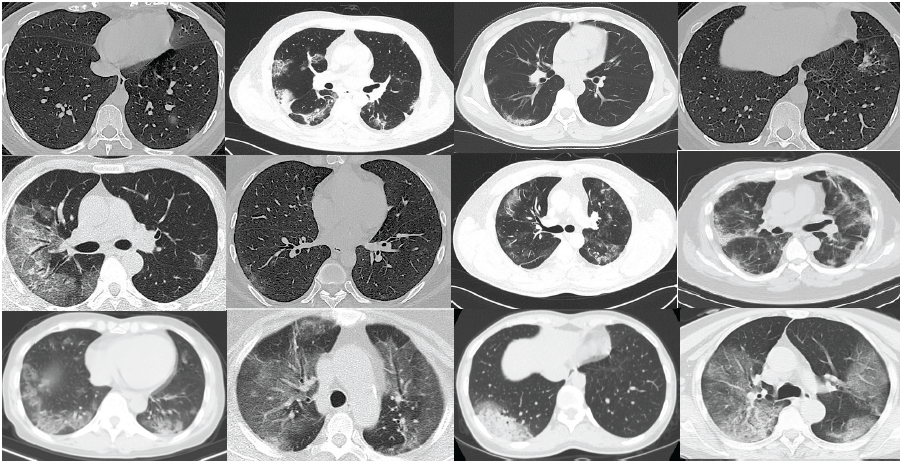
Examples of CT scans that are positive for COVID-19.

Transfer learning aims to leverage data-rich source-tasks to help with the learning of a data-deficient target task (CT-based diagnosis of COVID-19 in our case). One commonly used strategy is to learn a powerful visual feature extraction deep network by pretraining this network on large datasets in the source tasks and then adapt this pretrained network to the target task by finetuning the network weights on the small-sized dataset in the target task. While effective in general, transfer learning may be suboptimal due to the fact that the source data may have a large discrepancy with the target data in terms of visual appearance of images and class labels, which causes the feature extraction network biased to the source data and generalizes less well on the target data. We design different transferring strategies and perform a comprehensive study in the dimensions of source-target domain difference and neural architectures to investigate the effects of transfer learning for COVID-19 diagnosis and provide insightful findings.

Based on these findings, we propose **Self-Trans**, a self-supervised transfer learning approach where contrastive self-supervised learning [3] is integrated into the transfer learning process to adjust the network weights pretrained on source data, so that the bias incurred by source data is reduced. In self-supervised learning (SSL), we construct auxiliary tasks on CT images where the supervised labels in these tasks are solely from the images themselves without using any human annotations. Then we adjust the network weights by solving these auxiliary tasks. In these auxiliary tasks, the input images are in the same domain as the data in the target task and no human-annotated labels are used. Therefore, the bias to source images and their class labels can be effectively reduced.

### A. Contributions

Contributions of this paper are summarized as follows:

- We propose a sample-efficient deep learning system to facilitate the diagnosis of COVID-19 based on CT scans. Code will be open-sourced.
- To train and evaluate the system, we collect the COVID19-CT dataset^2^, which contains 349 positive CT scans with clinical findings of COVID-19, and 397 negative images without findings of COVID-19. To the best of our knowledge, this is the largest publicly-available CT dataset for COVID-19.
- We design different transferring strategies and perform a comprehensive study to investigate the effects of transfer learning for COVID-19 diagnosis and provide insightful findings.
- To learn from limited labeled data, we propose Self-Trans networks, which synergistically integrate contrastive self-supervised learning with transfer learning to learn powerful and unbiased feature representations for reducing the risk of overfitting.
- We perform extensive experiments to demonstrate the effectiveness of our proposed methods. It achieves an F1 score of 0.85, an AUC of 0.94, and an accuracy of 0.86 on the COVID19-CT dataset.

The rest of the paper is organized as follows. Section 2 reviews related works. Section 3 and 4 present the dataset, methods, and experiments. Section 5 concludes the paper.

## II. Related work

### A. Deep learning based diagnosis of COVID-19

Since the outbreak of COVID-19, there have been increasing efforts on developing deep learning methods to perform screening of COVID-19 based on medical images such as CT scans and chest X-rays. Wu et al. established an early-screening model based on multiple CNN models to classify CT scans of patients with COVID-19 [4]. Wang et al. proposed a 3D deep CNN (DeCoVNet) to detect COVID-19 [5] using chest CT slices. Chowdhury et al. employed CNN to identify COVID-19 patients based on chest x-ray images [6]. Several works have also applied 3D deep learning models to screen COVID-19 based on chest CT images [7], [8]. Yang et al. developed a deep learning based CT diagnosis system (DeepPneumonia) to assist clinicians to identify patients with COVID-19 [9]. Xu et al. developed a deep learning algorithm by modifying the inception transfer-learning model to provide clinical diagnosis ahead of the pathogenic test [10]. Shi et al. employed the “VB-Net” neural network to segment COVID-19 infection regions in CT scans [11]. Yu et al. constructed a system based on UNet++ for identification of COVID-19 from CT images [12]. Shen et al. proposed an infection-size-aware Random Forest (iSARF) method which can automatically categorize subjects into groups with different ranges of infected lesion sizes [13].

### B. Datasets about COVID-19

At present, few large-sized datasets with medical images on COVID-19 are publicly available due to privacy concerns and information blockade [14]. Existing datasets on COVID19 are mainly X-ray images [6], [14], [15]. The Italian Society of Medical and Interventional Radiology (SIRM) provides chest X-rays and CT images of 68 Italian COVID-19 cases [16]. Moore et al. released a dataset of axial and coronal CTs from 59 COVID-19 cases at Radiopaedia [17]. Other data sources provide medical images of no more than 10 patients [18], [19]. To deal with the lack of large-sized and open-source datasets containing CT images of COVID-19 cases, we built the COVID19-CT dataset by collecting medical images from COVID-19 related medRxiv and bioRxiv papers. Our dataset contains 349 COVID-19 positive CT scans from 216 COVID-19 cases. To our best knowledge, it is the largest public COVID-19 CT collection to date.

### C. Transfer learning

Transfer learning is normally performed by taking a standard neural architecture along with its pretrained weights on large-scale datasets such as ImageNet [20], and then finetuning the weights on the target task. This idea has been successfully applied to visual recognition [21] as well as language comprehension [22]. In the medical domain, transfer learning has also been widely used in medical image classification and recognition tasks, such as tumor classification [23], retinal diseases diagnosis [24], pneumonia detection [25], and skin lesion and cancer classification [26], [27]. A recent study in [28] explores the properties of transfer learning for medical imaging tasks and finds that the standard large networks pretrained on ImageNet are often over-parameterized and may not be the optimal solution for medical image diagnosis. In this paper, we continue to investigate different strategies of transfer learning and integrate contrastive self-supervised learning into the transfer learning process to learn powerful and unbiased feature representations for reducing the risk of overfitting.

### D. Self-supervised learning

Self-supervised learning (SSL) aims to learn meaningful representations of input data without using human annotations. It creates auxiliary tasks solely using the input data and forces deep networks to learn highly-effective latent features by solving these auxiliary tasks. Various strategies have been proposed to construct auxiliary tasks, based on temporal correspondence [29], [30], cross-modal consistency [31], etc. Examples of auxiliary tasks include rotation prediction [32], image inpainting [33], automatic colorization [34], context prediction [35], etc. Some recent works study self-supervised representation learning based on instance discrimination [36] with contrastive learning. Oord et al. propose contrastive predictive coding (CPC) to extract useful representations from high-dimensional data [37]. Bachman et al. propose a self-supervised representation learning approach based on maximizing mutual information between features extracted from multiple views of a shared context [38]. Most recently, Chen et al. present a simple framework for contrastive learning (SimCLR) [39] with larger batch sizes and extensive data augmentation [40], which achieves results that are comparable with supervised learning. Momentum Contrast (MoCo) [41],[42] expands the idea of contrastive learning with an additional dictionary and a momentum encoder. While previous methods concentrate on utilizing self-supervision to learn universal representations regardless of labels, our Self-Trans instead aims to boost the performance of supervised transfer learning with self-supervised pretraining on unlabeled data. Inspired by [42], [43], we aim to leverage self-supervised learning for COVID-CT recognition for which there are limited COVID-19 samples but abundant unlabeled CTs.

## III. Methods

The lack of annotated CT scans about COVID-19 brings significant challenges for deep-learning-based diagnosis of COVID-19 using CT images. To address this problem, we build COVID19-CT, a dataset containing hundreds of CT images positive for COVID-19. Though the largest of its kind, COVID19-CT is still small, making deep learning models trained on this dataset prone to overfitting. To address this problem, we systematically investigate different transfer learning strategies and propose a new approach called Self-Trans, which synergistically integrates unsupervised in-domain self-supervised learning with supervised out-of-domain transfer learning to learn effective and unbiased visual feature representations that are robust to overfitting. In the following subsections, we first introduce the COVID19-CT dataset that we built, then present transfer learning and the proposed Self-Trans approach for sample-efficient diagnosis of COVID-19 from CT scans.

### A. COVID19-CT dataset

CT scans are promising in providing accurate, fast, and cheap screening and testing of COVID-19 [44]. To facilitate the diagnosis of COVID-19, we first build a publicly available COVID19-CT dataset, containing 349 CT scans that are positive for COVID-19 and 397 negative CT scans that are normal or contain other types of diseases. To build this dataset, we first collected 760 preprints about COVID-19 from medRxiv^3^ and bioRxiv^4^, posted from Jan 19^th^ to Mar 25^th^. Many of these preprints report patient cases of COVID-19 and some of them show CT scans in the reports. CT scans are associated with captions describing the clinical findings in the CTs. We used PyMuPDF^5^ to extract the low-level structure information from the PDF files of preprints and located all the embedded figures. The quality (including resolution, size, etc.) of figures is well-preserved. From the structure information, we identified the captions associated with figures. Given these extracted figures and captions, we first manually selected all CT scans. Then for each CT scan, we read the associated caption to judge whether it is positive for COVID-19. If not able to judge from the caption, we located the text analyzing this figure in the preprint to make a final decision. For any figure that contains multiple CT scans as sub-figures, we manually split it into individual CTs, as shown in Figure 2.

**Fig. 2.**
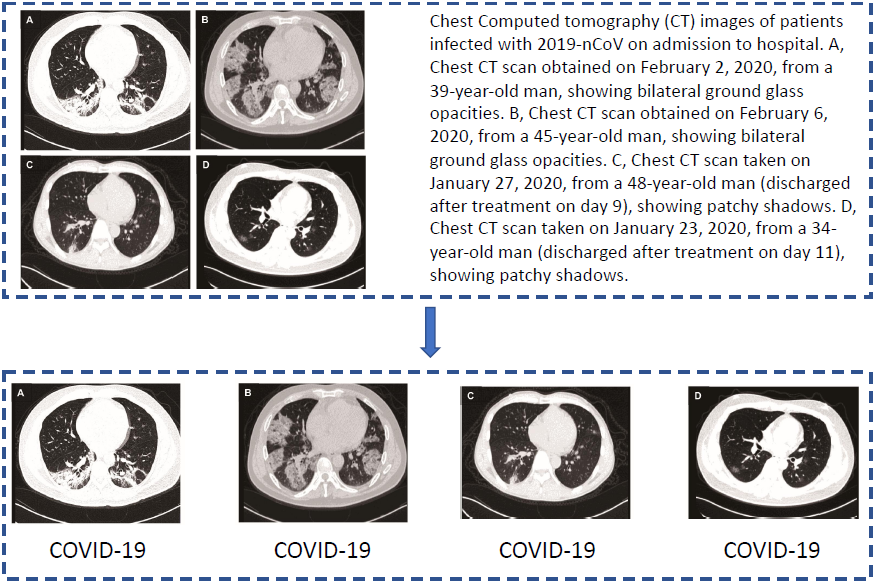
When building the COVID19-CT dataset, for any figure that contains multiple CT scans as sub-figures, we manually split it into individual CTs

In the end, we obtain 349 CT scans labeled as being positive for COVID-19. These scans are from 143 patient cases. The average, maximum, and minimum number of CT scans that a patient has is 1.6, 16.0, and 1.0 respectively. These CT images have different sizes. The average, maximum and minimum height are 491, 1853, and 153 respectively. The average, maximum, and minimum width are 383, 1485, and 124 respectively. Figure 1 shows some examples of the COVID-19 CT scans.

In addition to these COVID19-positive CTs, we also add 397 CTs that are negative for COVID-19. Among them, 202 negative CTs are selected from the PubMed Central (PMC)^6^ search engine. The rest 195 come from MedPix^7^, which is a publicly-open online medical image database that contains CT scans with various diseases.

### B. Transfer learning for CT-based COVID-19 diagnosis

Given a target task (e.g., diagnosing COVID-19 from CT scans in our case) that has limited training data, transfer learning aims to leverage large-scale data and human-provided labels from other source tasks to learn expressive and generalizable feature representations to help with the learning of the target task. A commonly used approach [45] is to pretrain a deep neural network — which is used for feature extraction — on large datasets in the source tasks by fitting the human-annotated labels therein, then fine-tune this pretrained network on the target task. In our case, we can take a classic neural architecture such as ResNet [46] and its weights pretrained on large-scale image classification datasets such as ImageNet, then fine-tune it on the COVID19-CT dataset, with the goal of transferring the images and classes labels in ImageNet into our task for mitigate the deficiency of COVID-19 CTs. When applying this strategy, we should keep several caveats in mind. First, the image data in source tasks have a large domain discrepancy with COVID-19 CTs. For example, the ImageNet images mostly belong to categories in the general domain, such as cat, dog, chair, etc. whereas the images in our task are CTs. The visual appearance, size, resolution of ImageNet images are quite different from chest CTs. As a result, the visual representations learned on ImageNet may not be able to represent CT images well, which casts doubts on the transferability from other sources of images to COVID-19 CTs. Second, the transferability across tasks depends on the neural architectures used for representation learning. Certain architectures facilitate transfer learning better than others.

In this work, we aim to perform a systematic study on how these factors affect the transferability from other image classification tasks to our task, and accordingly based on the study result, we design the optimal transfer learning strategy. To study the first factor — domain difference in data, we perform transfer learning on two datasets: one is ImageNet in the general domain; the other is the Lung Nodule Maligancy (LNM) dataset^8^ in the CT domain. Compared with ImageNet, LNM has a smaller domain discrepancy with our dataset, but has a smaller number of images and the images are less diverse. To study the second factor — neural architectures, we experiment different backbone networks, including VGG16 [47], ResNet18 [46], ResNet50 [46], DenseNet-121 [48], DenseNet-169 [48], EfficientNet-b0 [49], and EfficientNet-b1 [49]. We evaluate the efficacy of different transfer learning strategies with different pretrained datasets and network architectures via extensive experiments and provide insightful findings.

Considering that our dataset is small and our task is binary classification, to study whether there is indeed over-parameterization in the traditional ImageNet models when applied to COVID-19 diagnosis, in addition to these large-sized architectures, we also design a light-weight architecture as shown in Figure 3. The basic building block for this network is the combination of a 2d-convolution with an ReLU activation. The block repeats four times. There is an additional batch normalization layer in the first block and an average pooling layer after the third block. We refer to this network as CRNet, whose structure is as follows: (conv32-bn-relu), maxpool, (conv64-relu), maxpool, (conv128-relu), maxpool, global avgpool, and classification layer. The conv(n) represents a 2d convolutional layer with *n* output channels with a kernel size of 7*×*7 and a stride of 1. The bn denotes a 2d batch normalization [50] layer. The relu stands for an ReLU layer. The maxpool stands for max pooling with a kernel size of 3*×*3 and a stride of 2. The global avgpool is an average pooling layer with a kernel size of 2*×*2 and a stride of 2.

**Fig. 3.**
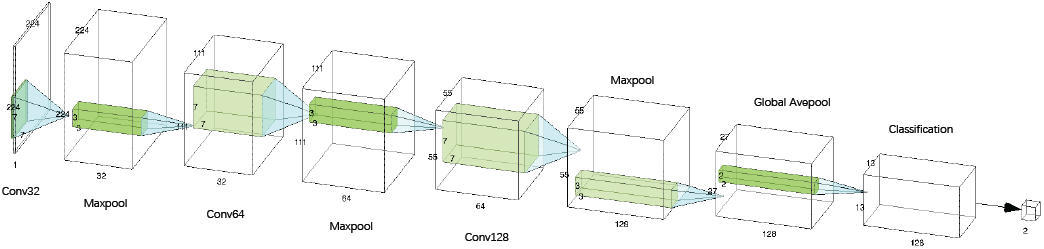
CRNet architecture. Please zoom to view better

### C. Contrastive self-supervised learning for CT-based COVID-19 diagnosis

As discussed in the above section, while the popular transfer learning methods, which pretrain the model on large-scale datasets and fine-tune it on the target dataset, are helpful in improving the performance on the target task, there are two concerns about the transferability of the source data. First, the source data has a large domain shift from the target data. For example, the images in ImageNet are mostly in the natural image domain while the images in COVID-19 diagnosis tasks are from the medical domain. Second, in transfer learning, pretraining is conducted by fitting the class labels in the source domain and these classes are largely different from those in the target task. For example, the classes in ImageNet are mostly about non-medical concepts such as dog, cat, desk, etc. while the labels in our target task are COVID and Non-COVID. Trying to fit the natural-domain class labels may cause the representations learned in the pretraining to be biased to these natural classes and less-well generalized to the COVID CTs.

To further solve this problem, we propose an **Self-Trans** approach, which integrates contrastive self-supervision [3] into the transfer learning process. Self-supervised learning (SSL) [36], [39], [41], [42] is a learning paradigm which aims to capture the intrinsic patterns and properties of input data (e.g., CT images) without using human-provided labels. The basic idea of SSL is to construct some auxiliary tasks solely based on the data itself without using human-annotated labels and force the network to learn meaningful representations by performing the auxiliary tasks well. Typical self-supervised learning approaches generally involve two aspects: constructing auxiliary tasks and defining loss functions. The auxiliary tasks are designed to encourage the model to learn meaningful representations of input data without utilizing human annotations. The loss functions are defined to measure the difference between a model’s prediction and a fixed target, the similarities of sample pairs in a representation space (e.g., contrastive loss), or the difference between probability distributions (e.g., adversarial loss). In this work, we design the auxiliary tasks based on the contrastive loss [41], [42], [51] to provide self-supervision for the transfer learning process. To be specific, the auxiliary task is to judge whether two images created via random data augmentation are augments of the same original image. We build a large and consistent dictionary on-the-fly based on the contrastive loss to fulfill this auxiliary task. To fully explore the structure and information of the CT images, we apply Self-Trans on both external large-scale lung CT datasets and our collected COVID19-CT dataset.

#### 1) Contrastive learning for self-supervision

Given an original image in the dataset, contrastive self-supervised learning (CSSL) [51] performs data augmentation of this image and obtains two augmented images: ***x***_*q*_ and ***x***_*k*_, where the first one is referred to as query and the second one as key. Two networks *f*_*q*_(; *θ*_*q*_) and *f*_*k*_(; *θ*_*k*_), referred to as the query encoder and the key encoder and parameterized by weights *θ*_*q*_ and *θ*_*k*_, are used to obtain latent representations — ***q*** = *f*_*q*_(***x***_*q*_; *θ*_*q*_) and ***k*** = *f*_*k*_(***x***_*k*_; *θ*_*k*_) — of the query and key images respectively. A query and a key belonging to the same image are labeled as a positive pair. A query and a key belonging to different images are labeled as a negative pair. The auxiliary task is: given a (query, key) pair, judging whether it is positive or negative.

Implementation-wise, CSSL uses a queue to store a set of keys ***k***_*i*_ from different images. Given a new pair (***q***_*j*_, ***k***_*j*_) obtained from a new image, a contrastive loss can be defined as:

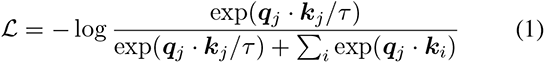

where *τ* is an annealing parameter. The weights in the encoders are learned by minimizing losses of such a form.

#### 2) Momentum encoder with queue-structured dictionary

Existing methods adopt various mechanisms to preserve and sample key vectors [37], [39], [51], [52]. A **Siamese-like** solution is to use the same network *f*_*k*_ = *f*_*q*_ on ***x***_*k*_ and ***x***_*f*_ simultaneously. Extreme large mini-batch (batch-size up to 8192 [39]) is required to learn discriminative features from contrasting. This method is straightforward but incredibly expensive in terms of computational resources. Another option is to store the representations of historical keys in a negative key dictionary *D*_*k*_ = {***k***_*i*_*}*, called **memory bank** [36]. At each iteration, a mini-batch of keys are sampled from the memory bank instead of using *f*_*k*_. The current mini-batch of queries is updated to the memory bank for replacement. This design inherently gets rid of large batch-size with an extended buffer pool. However, the key sampling step involves inconsistency for training the encoder.

Momentum Contrastive (MoCo) [41] learning tolerates and integrates from both. A queue-structured key dictionary with fixed length replaces the memory bank. According to the first-in-first-out (FIFO) nature of the queue, the oldest key mini-batch will serve as the negative keys and be replaced by the new queries. This mechanism prevents irregular sampling of negative sampling.

Another essential component of this model is that, we neither update the key encoder with back-propagation nor blindly copy the query encoder to the key encoder, but keep running average of the key encoder *f*_*k*_ [41], [53]. It can also be called a *momentum encoder*. The updating rule of *θ*_*k*_ and *θ*_*q*_ can be formulated as

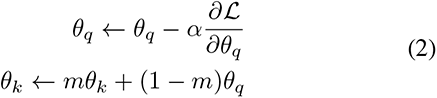

In the equation, *m* = 0.999 is the momentum coefficient and *α* is the learning rate of query encoder. Only *θ*_*q*_ is updated through back-propagation, whereas *θ*_*k*_ maintains a weighted average of the past states. As suggested in Figure 4, we adopt the self-supervised learning prior to supervised training on the COVID-CT data, as a kind of weight initialization. This mechanism has proved in experiments that it can further improve the model’s performance in our CT classification task. The detailed algorithm of Self-Trans is shown in Algorithm 1.

**Fig. 4.**
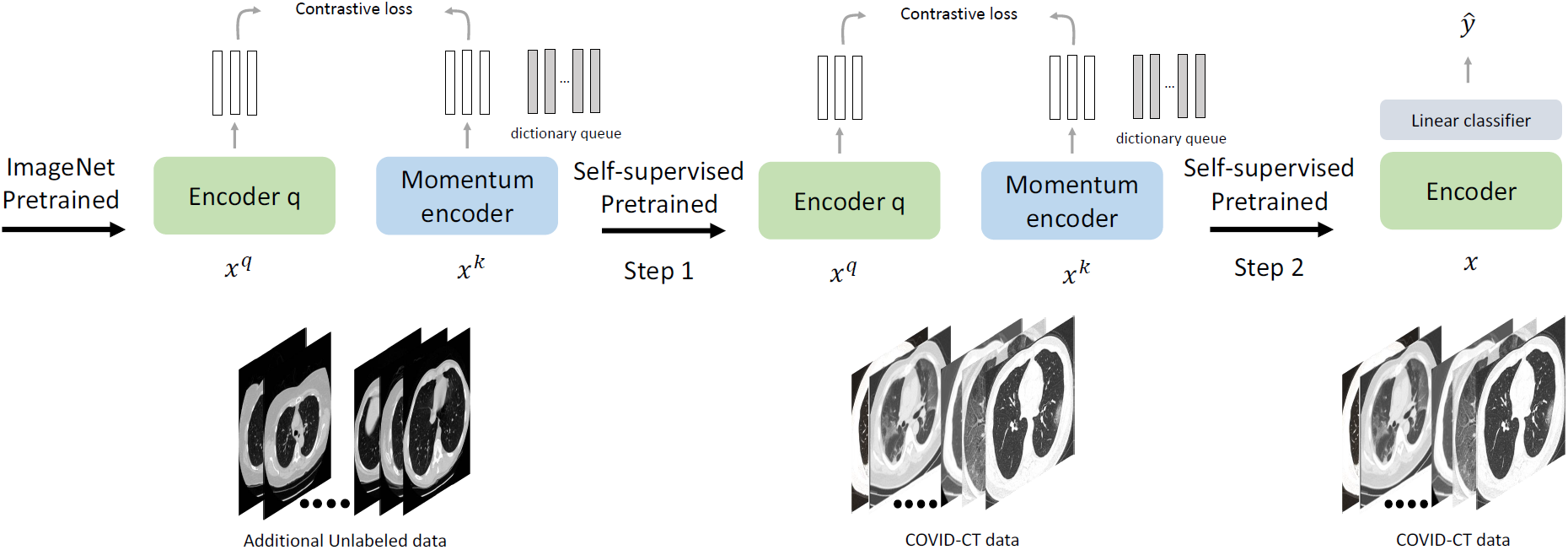
Framework and training pipeline of Self-Trans. Please zoom to view better

## IV. Experiments

To demonstrate the efficacy of our proposed approach and investigate the effects of transfer learning and self-supervised learning, we extensively evaluate the randomly initialized networks, ImageNet transferred networks, and our proposed Self-Trans networks. In the following subsections, we will introduce the datasets, experimental settings, and results for these three series of approaches. Please note that the ablation study is included in the Results subsection for each series of approaches.

### A. Datasets

Our collected COVID19-CT dataset consists of 349 COVID-19 CTs and 397 Non-COVID-19 CTs. The CT images were resized to 224 × 224. We split the dataset into a training set, a validation set, and a test set by patient IDs with a ratio of 0.6: 0.15: 0.25. Table I shows the statistics of the three sets. In addition to our COVID-19 CT dataset, we also include images from the Lung Nodule Analysis (LUNA) [54] database as a source of additional unlabeled CT data. It is originally designed for lung nodule detection and segmentation. From the total 888 CT scans, we randomly select 500 subjects and from each we extract two CT slices that contain annotated lesions or lung shadow. These images are not included in the COVID19-CT dataset as negative examples but are served as unlabelled images for self-supervised learning.

**TABLE I.**
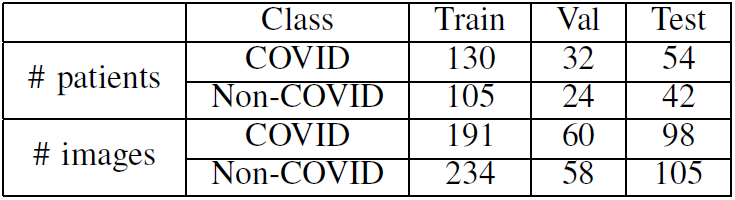
Dataset Split

|  | Class | Train | Val | Test |
| --- | --- | --- | --- | --- |
| # patients | COVID | 130 | 32 | 54 |
|  | Non-COVID | 105 | 24 | 42 |
| # images | COVID | 191 | 60 | 98 |
|  | Non-COVID | 234 | 58 | 105 |

#### Algorithm 1 Algorithm of Self-Trans

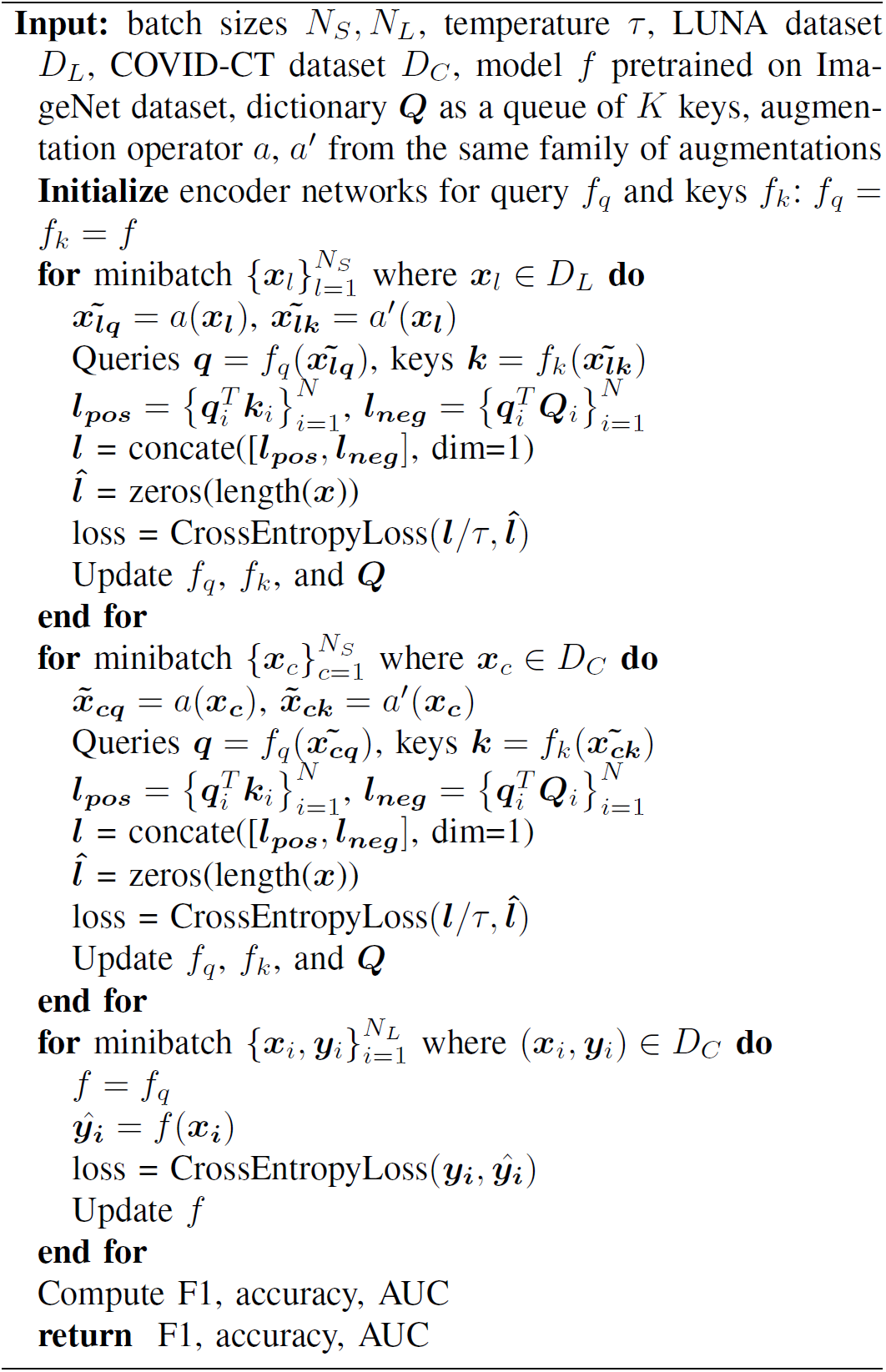

### B. Experimental Settings

The models are implemented in PyTorch. Batch normalization [55] is used through all models. Binary cross-entropy serves as the loss function. The networks are trained with four GTX 1080Ti GPUs using data parallelism. Hyperparameters are tuned on the validation set. Data augmentation is implemented to further improve generalization. For each image in our COVID19-CT dataset, we apply different random affine transformations including random cropping with a scale of 0.5 and horizontal flip. Color jittering is also applied with random contrast and random brightness with a factor of 0.2.

We evaluate our approaches using five metrics: (1) Accuracy, which measures the percentage of diagnostic predictions that match exactly with the ground-truth; (2) Precision, which is the fraction of true positives among the predicted positives; (3) Recall, which is the fraction of the total number of true positives that are predicted as positive; (4) F1-score, which is the harmonic mean of precision and recall; (5) AUC, which is the area under the receiver operating characteristic curve showing how false positive rate increases as true positive rate increases. For all five metrics, the higher, the better.

### C. Evaluations on Randomly Initialized Networks

To demonstrate the efficacy of our proposed approach and investigate the effects of transfer learning and self-supervised learning, we first experiment on the randomly initialized networks with different backbones as baselines. The backbone networks include VGG-16 [47], ResNet-18 [46], ResNet-50 [46], DenseNet-121 [48], DenseNet-169 [48], EfficientNet-b0 [49], EfficientNet-b1 [49], and our proposed CRNet.

#### 1) Implementation Details

For classifiers trained from scratch, the Adam [56] optimizer is used with an initial learning rate of 0.0001 and a mini-batch size of 16. The cosine annealing scheduler is applied on the optimizer with a period of 10 to adjust the learning rate across the training process. We train our models with 50 epochs. We initialize the weights with Kaiming Initialization [57].

#### 2) Results

Table III shows the evaluation results (columns marked with “Rand.”) for neural networks trained with random initialization. Comparing ResNet-18 with ResNet-50 and comparing DenseNet-121 with DenseNet-169, we can see that deeper networks generally yield higher classification performance. The performance also benefits a lot from more sophisticated network structure like residual connection [46] and dense connection [48].

Table II shows the number of weight parameters in different networks. The CRNet has a much smaller number of parameters but the performance is on par with or better than more sophisticated architectures such as ResNet-50.

**TABLE II.** Comparison of parameter numbers

|  | Parameter numbers |
| --- | --- |
| VGG-16 | 138,357,544 |
| ResNet-18 | 11,689,512 |
| ResNet-50 | 25,557,032 |
| DenseNet-121 | 7,978,856 |
| DenseNet-169 | 14,149,480 |
| EfficientNet-b0 | 5,288,548 |
| EfficientNet-b1 | 7,794,184 |
| CRNet | 546,882 |

### D. Evaluations on Large-Scale Dataset Transferred Networks

After evaluating the randomly-initialized networks, we thoroughly investigate the performance of networks pretrained on large-scale datasets, including ImageNet and the Lung Nodule Maligancy (LNM) dataset, with different backbones, including VGG-16, ResNet-18, ResNet-50, DenseNet-121, DenseNet-169, EfficientNet-b0, EfficientNet-b1, and CRNet.

The results of transfer learning from ImageNet are shown in Table III (columns marked with “Trans.”), where we first train the networks from scratch on ImageNet and then fine-tune them on the COVID19-CT dataset. Comparing these results with those achieved by randomly initialized networks (columns marked with “Rand.” in Table III), we can see pretraining on ImageNet significantly improves classification performance. This demonstrates the effectiveness of transfer learning, which leverages large-scale images and their class labels in source tasks to help with the learning of the target task. In certain cases, the benefits of transfer learning are highly significant. For example, for VGG16, when trained with random initialization, it performs the poorest. Transfer learning helps it to improve accuracy by 10% (absolute improvement), F1 by 8% (absolute), and AUC by 8% (absolute), exceeding the performance of networks (e.g., ResNet) that have more sophisticated architectures designed for preventing overfitting, even when these networks are pretrained as well.

**TABLE III.** Performance comparison between randomly initialized networks (Rand.) and ImageNet pretrained networks (Trans.)

|  | Accuracy |  | F1 |  | AUC |  |
| --- | --- | --- | --- | --- | --- | --- |
|  | Rand. | Trans. | Rand. | Trans. | Rand. | Trans. |
| VGG-16 | 0.66 | 0.76 | 0.58 | 0.76 | 0.74 | 0.82 |
| ResNet-18 | 0.67 | 0.74 | 0.66 | 0.73 | 0.76 | 0.82 |
| ResNet-50 | 0.69 | 0.80 | 0.72 | **0.81** | 0.76 | 0.88 |
| DenseNet-121 | 0.76 | 0.79 | 0.77 | 0.79 | 0.82 | 0.88 |
| DenseNet-169 | 0.80 | **0.83** | 0.79 | **0.81** | 0.86 | 0.87 |
| EfficientNet-b0 | 0.72 | 0.77 | 0.71 | 0.78 | 0.76 | **0.89** |
| EfficientNet-b1 | 0.70 | 0.79 | 0.62 | 0.79 | 0.77 | 0.84 |
| CRNet | 0.72 | 0.73 | 0.76 | 0.76 | 0.77 | 0.79 |

We observe that for CRNet which is a small-sized network, the effect of transfer learning is marginal. This is probably because a small-sized network is more robust to overfitting, therefore it has a smaller need of using transfer learning for combating overfitting. On the contrary, for large-sized neural architectures such as VGG-16, initialization with pretrained weights makes a huge difference. The reason is that large-sized networks are more prone to overfitting, especially considering that our dataset is fairly small. Under such circumstances, transfer learning has a better chance to play its value.

Table IV shows the performance of the DenseNet-169 backbone with weights (1) randomly initialized, (2) pretrained on ImageNet, (3) pretrained on LMN, and (4) pretrained first on ImageNet, then on LMN. From this table, we make the following observations. First, transfer learning on either ImageNet or LMN improves performance, which further demonstrates the efficacy of transfer learning. Second, the performance of the network pretrained on ImageNet has no significant difference with that pretrained on LMN (the former has slightly better accuracy but worse F1). The two datasets have both advantages and disadvantages. ImageNet has more images and more classes than LMN, which enables learning more powerful and generalizable feature representations. The downside of ImageNet is that its images have a large domain discrepancy with the CTs in COVID19-CT whereas the images in LMN are all CTs. The advantages and disadvantages of these two datasets make them similar in providing transfer learning values. Pretraining first on ImageNet then on LMN achieves better performance than just pretraining on ImageNet. This shows that using data with complementary properties (e.g., size, domain similarity) can generate a synergistic effect in transfer learning.

**TABLE IV.** Performance of DenseNet-169 with different weights initialization mechanisms

|  | Accuracy | F1 | AUC |
| --- | --- | --- | --- |
| Random Initialization | 0.80 | 0.79 | 0.86 |
| Pretrain on ImageNet | **0.83** | 0.81 | 0.87 |
| Pretrain on LMN | 0.80 | 0.82 | 0.87 |
| Pretrain first on ImageNet, then on LMN | 0.81 | **0.82** | **0.89** |

### E. Evaluation of Self-Trans

In this section, we evaluate the performance of our proposed Self-Trans networks, and compare them with networks pre-trained on large-scale datasets. Given the weights pretrained on other datasets, we leverage contrastive self-supervised learning (CSSL) to further train these weights on CT images in our COVID19-CT dataset. Note that in this step, the labels of these CT images are not utilized. CSSL is only performed on the CTs themselves. After CSSL training, we fine-tune the weights on both the CTs and the class labels by optimizing the classification loss.

#### 1) Self-supervised baselines

To address the effectiveness of self-supervision, we also establish a baseline model with self-supervised auxiliary task. In this paper, we select the image rotation prediction [32] as auxiliary task in multi-task learning scheme. For each training image ***x***, we randomly rotate it with angle *ϕ* ∈ {0°, 90°, 180°, 270°}. A 4-way rotation prediction classifier and CT classification classifier share the same feature extractor. Losses for both tasks are added together and model is jointly trained. We do not rotate samples at test time.

#### 2) Additional Experimental Settings

Following the same setting in MoCo, we added a 2-layer multi-layer perceptron (MLP) head with 2048 hidden units. The size of the dynamic dictionary was set to 512. Stochastic gradient descent (SGD) was used as the optimizer for self-supervised learning (SSL), with a minibatch size of 128, a weight decay of 0.0001, a momentum of 0.9, and an initial learning rate of 0.015. The learning rate was adjusted by the cosine learning rate scheduler. The training was conducted on 4 GPUs with data parallelism. We carefully design data augmentation methods to serve as the pretext tasks for the Self-Trans methods. Specifically, we utilize data augmentation including random horizontal flip, random cropping with a size of 0.2 in area, random color jittering such as random brightness with a ratio of 0.4, random contrast of 0.4, random saturation of 0.4, random hue of 0.1, Gaussian blur, and random gray-scale conversion.

#### 3) Results

Table V shows the results of Self-Trans when applied to ResNet-50 and DenseNet-169. As can be seen from this table, Self-Trans achieves much better performance than vanilla transfer learning (Trans). The possible reason is that the weights learned by transfer learning may be biased to the images and class labels in the source tasks and generalize less well on the target task. SSL can help to reduce this bias by adjusting the weights using the data of the target task and without using any labels.

**TABLE V.** Comparison between Self-Trans and vanilla transfer learning (Trans)

|  | Accuracy | F1 | AUC |
| --- | --- | --- | --- |
| ResNet-50 (Trans) | 0.80 | 0.81 | 0.88 |
| ResNet-50 (Self-Trans) | 0.84 | 0.83 | 0.91 |
| DenseNet-169 (Trans) | 0.83 | 0.81 | 0.87 |
| DenseNet-169 (Self-Trans) | **0.86** | **0.85** | **0.94** |

#### 4) Ablation Studies

To fully understand the effects of self-supervised learning, we conduct the following ablation studies.

- **Method 1**: Randomly initialize weights. Perform SSL on the COVID19-CT dataset without using COVID/Non-COVID labels. Then fine-tune on COVID19-CT using labels.
- **Method 2**: Pretrain on ImageNet. Perform SSL on COVID19-CT without using labels and with pretrained weights. Then fine-tune on COVID19-CT using labels.
- **Method 3**: Pretrain on ImageNet. Perform SSL on the LUNA dataset without using labels of LUNA. Then fine-tune on COVID19-CT using labels.
- **Method 4**: Pretrain on ImageNet. Perform the auxiliary task of rotation predication as SSL baseline. Jointly learn rotation prediction and COVID19-CT classification.
- **Self-Trans**: Pretrain on ImageNet. Perform SSL on LUNA without using labels of LUNA. Then perform SSL on COVID-CT without using labels of COVID-CT, and finally fine-tune on COVID19-CT using labels.

Table VI shows the results in different ablation study settings, conducted on ResNet-50 and DenseNet-169 backbones. From this table, we observe the following. First, Method 2 which performs SSL on top of transfer learning works much better than Method 1 which performs SSL without using transfer learning. This demonstrates that it is more effective to apply SSL on pretrained weights instead of from scratch. Second, Method 2 which performs SSL on COVID19-CT (without using labels) largely outperforms Method 3 which performs SSL on LUNA. This implies that it is more effective to apply SSL directly on the data in the target task than on external data. Third, comparing Method 3 and Self-Trans, it is further confirmed that performing SSL directly on the data of the target task achieves better performance (by Self-Trans). Forth, Method 4 with an SSL auxiliary task beats the vanilla transfer learning counterpart, but do not surpass the SSL model with contrastive learning in Method 2 and Self-Trans. Such experimental results not only illustrate the effectiveness of SSL, but also provide concrete evidence that CSSL has stronger feature representation learning capabilities than traditional SSL methods. Fifth, Self-Trans performs slightly better than Method 2, which demonstrates that performing SSL on external data is also helpful, though not as useful as performing SSL directly on target-task’s data.

**TABLE VI.** Performance of Self-Trans in the ablation studies

|  | Accuracy | F1 | AUC |
| --- | --- | --- | --- |
| ResNet-50 (Method 1) | 0.69 | 0.63 | 0.66 |
| ResNet-50 (Method 2) | 0.83 | 0.83 | 0.91 |
| ResNet-50 (Method 3) | 0.75 | 0.78 | 0.85 |
| ResNet-50 (Method 4) | 0.81 | 0.81 | 0.88 |
| ResNet-50 (Self-Trans) | 0.84 | 0.83 | 0.91 |
| DenseNet-169 (Method 1) | 0.70 | 0.63 | 0.77 |
| DenseNet-169 (Method 2) | 0.86 | 0.85 | 0.91 |
| DenseNet-169 (Method 3) | 0.74 | 0.76 | 0.86 |
| DenseNet-169 (Method 4) | 0.83 | 0.82 | 0.88 |
| DenseNet-169 (Self-Trans) | **0.86** | **0.85** | **0.94** |

Another thing that we are interested in investigating is: given the weights of the feature extractor learned by SSL, when fine-tuning the overall classification network on the COVID19-CT images and labels, should we just fine-tune the final classifier layer or fine-tune the weights of the feature extractor as well? Table VII shows the results on two backbones: ResNet-50 and DenseNet-169, where “frozen” denotes that the weights of the feature extractor are not fine-tuned during the fine-tuning process and “unfrozen” denotes that these weights are fine-tuned together with those in the final classification layer. As can be seen, fine-tuning feature extraction weights yields much better performance. This is because using class labels to fine-tune these weights can make the extracted features more discriminative and hence more effective in distinguishing COVID-19 CTs from Non-COVID-19 CTs.

**TABLE VII.** Performance with the weights of the feature extractor frozen and unfrozen

|  | Accuracy | F1 | AUC |
| --- | --- | --- | --- |
| ResNet-50 (Frozen) | 0.70 | 0.63 | 0.77 |
| ResNet-50 (Unfrozen) | 0.83 | 0.83 | 0.91 |
| DenseNet-169 (Frozen) | 0.79 | 0.77 | 0.83 |
| DenseNet-169 (Unfrozen) | **0.86** | **0.85** | **0.91** |

Figure 5 shows the Grad-CAM [58] visualizations for DenseNet-169 trained from baseline methods and our proposed Self-Trans. By comparing Column (3) with Column (5), we notice that the DenseNet-169 model trained with random initialization erroneously focuses on some image edges and corners that are not related to COVID-19. In contrast, transfer learning methods generally lead to more accurate disease-related visual localization. By comparing Column (5) with Column (7), we can see that our proposed Self-Trans method can have even better localization of the disease region than the ImageNet pretrained method.

**Fig. 5.**
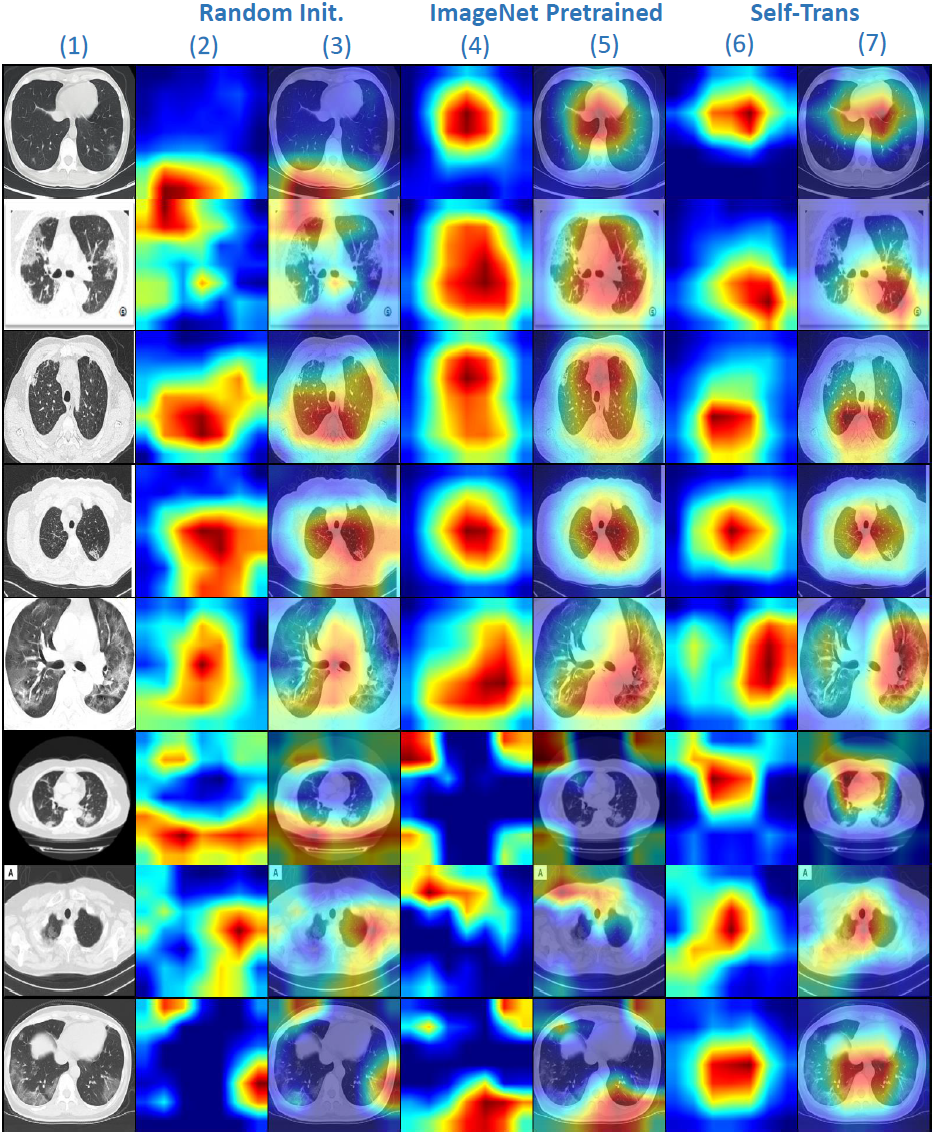
Grad-CAM visualizations for DenseNet-169. From left to right: Column (1) are original images with COVID-19; Column (2-3) are Grad-CAM visualizations for the model trained with random initialization; Column (4-5) are Grad-CAM visualizations for ImageNet pretrained model; Column (6-7) are Grad-CAM visualizations for Self-Trans model.

## V. Conclusions

In this paper, we study how to develop sample-efficient deep learning methods to accurately diagnose COVID-19 from CT scans. To facilitate the open research in this area, we build COVID19-CT, a dataset containing 349 CT scans positive for COVID-19. To our best knowledge, it is the largest COVID19-CT dataset that is publicly available to date. Though the largest, it still incurs a high risk of overfitting for data-hungry deep learning models. To reduce this risk, we develop data efficient methods that are able to mitigate data deficiency. We propose Self-Trans, a self-supervised transfer learning approach that learns expressive and unbiased visual feature representations that are robust to overfitting. Through extensive experiments, we demonstrate the effectiveness of our methods.

## Data Availability

The data is publicly available.

https://github.com/UCSD-AI4H/COVID-CT

## Footnotes

1 https://coronavirus.1point3acres.com/en/world

2 https://github.com/UCSD-AI4H/COVID-CT

3 https://www.medrxiv.org/

4 https://www.biorxiv.org/

5 https://github.com/pymupdf/PyMuPDF

6 https://www.ncbi.nlm.nih.gov/pmc/

7 https://medpix.nlm.nih.gov/home

8 https://www.kaggle.com/kmader/lungnodemalignancy/home

